# Public perceptions and interactions with UK COVID-19 Test, Trace and Isolate policies, and implications for pandemic infectious disease modelling

**DOI:** 10.1101/2022.01.31.22269871

**Authors:** Guy C. Marshall, Rigina Skeva, Caroline Jay, Miguel E. P. Silva, Martyn Fyles, Thomas House, Emma L. Davis, Li Pi, Graham F. Medley, Billy J. Quilty, Louise Dyson, Lucy Yardley, Elizabeth Fearon

**Affiliations:** Department of Computer Science, University of Manchester, Manchester M13 9PL, UK; Department of Mathematics, University of Manchester, Manchester M13 9PL, UK; The Alan Turing Institute, London, UK; Big Data Institute, Li Ka Shing Centre for Health Information and Discovery, University of Oxford, Oxford OX3 7LF, UK; Centre for the Mathematical Modelling of Infectious Disease, London School of Hygiene and Tropical Medicine (LSHTM), London WC1E 7HT, UK; Department of Global Health and Development, LSHTM, London WC1E 7HT, UK; Department of Infectious Disease Epidemiology, LSHTM, London WC1E 7HT, UK; The Zeeman Institute for Systems Biology & Infectious Disease Epidemiology Research, School of Life Sciences and Mathematics Institute, University of Warwick, Coventry CV4 7AL, UK; Health Protection Research Unit in Behavioural Science, University of Bristol, Bristol BS8 1QU, UK; School of Psychology, University of Southampton, Southampton SO17 1BJ, UK

## Abstract

The efforts to contain SARS-CoV-2 and reduce the impact of COVID-19 have been supported by Test, Trace and Isolate (TTI) systems in many settings, including the United Kingdom. The mathematical models underlying policy decisions about TTI make assumptions about behaviour in the context of a rapidly unfolding and changeable emergency. This study investigates the reported behaviours of UK citizens in July 2021, assesses them against how a set of TTI processes are conceptualised and represented in models and then interprets the findings with modellers who have been contributing evidence to TTI policy. We report on testing practices, including the uses of and trust in different types of testing, and the challenges of testing and isolating faced by different demographic groups. The study demonstrates the potential of input from members of the public to benefit the modelling process, from guiding the choice of research questions, influencing choice of model structure, informing parameter ranges and validating or challenging assumptions, to highlighting where model assumptions are reasonable or where their poor reflection of practice might lead to uninformative results. We conclude that deeper engagement with members of the public should be integrated at regular stages of public health intervention modelling.

## 1 Introduction

The COVID-19 pandemic, caused by infection with the SARS-CoV-2 virus, has resulted in the illness and deaths of many people in the UK and internationally. Damage to health and well-being has occurred both directly from the disease and indirectly via its disruptive effects on society and the economy. In the UK, the Test, Trace and Isolate system (TTI) has been used population-wide from May 2020 (with some differences across England and devolved nations), to help control transmission by asking people who have been identified as infected, or who have a high likelihood of having been infected, to isolate or quarantine within their households to prevent onward transmission in society. Early modelling work suggested that the characteristics of the virus, particularly the extent of pre-symptomatic and asymptomatic transmission, coupled with high overall infectiousness, would create major challenges for the use of TTI to control the pandemic and TTI would likely need to be implemented alongside other interventions to limit social contact. Performance of the TTI system would need to be high, with a high proportion of cases identified and isolation, with tracing and quarantine effectively implemented with minimal delays and a high level of coverage [48, 49, 50]. Achieving this depends not only on testing and tracing design and technologies, but also critically on the extent to which members of the public are informed and supported in engaging with each of the multiple TTI steps required. An understanding of the public’s engagement with TTI policies can in turn be used to model, evaluate and develop improved TTI protocols.

The specific details of TTI policies have changed over time, though some aspects such as the symptoms that prompt testing (new, continuous cough, fever and/or loss of smell or taste) and case isolation in the home among those testing positive, have remained unchanged. As of July 2020, when an individual experienced one of the above symptoms, they were asked to book and take a polymerase chain reaction (PCR) test at a testing site or ordered to the home, and were asked to isolate along with all their household members until receiving a negative result, which usually took 24-48 hours. All needed to remain ’isolated’ (cases) or ’quarantined’ (contacts of the case) at home until, and if, a negative result was received by phone or text message. If the PCR test result was positive, cases were interviewed to ascertain their close contacts in the period from 2 days prior to 7 days post becoming symptomatic and these individuals were traced and notified, while the case was asked to notify their household members. Household and non-household contacts were asked to quarantine at home for 10 days.

At the time of this study, July 2021, UK COVID-19 pandemic policy in general was rapidly changing. On 19th July, midway through the study, England’s laws related to infection prevention were relaxed, including requirements to wear masks, limitations on social contact and social distancing rules. The requirement to test and isolate if symptomatic was retained throughout this period, as was the requirement to quarantine if having contacted someone infected [1]. Rapid lateral flow device (LFD) tests were freely available in pharmacies, some workplaces and via a government website throughout the period, and PCR tests were freely available for those with the specific symptoms and for contacts of identified cases. Pandemics are evolving situations, and the qualitative snapshot we have taken complements previous TTI policy-related work of March-April 2020 [2], TTI pilots of daily testing in December 2020 [4], Sheffield-specific interviews from October 2020 - February 2021 [5] and the CORSAIR surveys of June 2021 [6], and Office for National Statistics (ONS) surveys (e.g. [47]).

This study was conducted as part of a 12 month ’rapid response’ academic TTI epidemiological modelling research project, which aimed to support the design and deployment of TTI policies, reporting findings via UK government advisory channels, primarily the Scientific Pandemic Influenza Group on Modelling (SPI-M) and the Scientific Advisory Group for Emergencies (SAGE) [46]. The majority of work undertaken in this project was responsive to requests via these advisory channels, to provide fast advice relating to specific policy decisions. The present study was undertaken towards the end of this project, with the aim to understand better how well the modelling fit to practices, and to capture information to inform ongoing TTI policy and modelling, and to inform how we can improve upon modelling of public health interventions for future pandemics. The high level research questions are:

- What are public attitudes towards TTI and how do members of the public interact with TTI tools and policies in practice?
- How might pandemic modellers reflect this information in their TTI intervention modelling?

We used thematic analysis to analyse our interview findings and then presented the results to pandemic response modellers in a workshop, asking them to reflect on how the findings aligned with assumptions about how TTI worked within their models, appraise how what they had learned about public interaction with TTI in practice might change their modelling approach, and to consider the implications for pandemic modelling going forward. We then invited workshop participants to co-write the relevant sections of this manuscript.

The paper is organised into sections. The Methods section describes the interviews and approach to engaging members of the public with how TTI processes were modelled, the analysis protocol, and engagement with modellers. The Results section describes the findings from our thematic analysis, and the Discussion section places the results in the context of prior studies on testing, contact tracing and isolation, and briefly explores implications for policy, while the “Reflections on implications for modelling” sub-section incorporates feed-back from TTI modellers about this study, and shares their thoughts on the impact of public involvement in informing modelling practices. The Conclusion section summarises learnings to take forward for the ongoing management of SARS CoV-2 but also practices for future pandemic planning.

## 2 Methods

### 2.1 Interview Study

#### 2.1.1 Participants and recruitment

Participants were recruited via purposive, snowballing and convenience sampling. The inclusion criteria for participating in the online interviews were (1) to be 18 years old or older and (2) live in the UK. Participants were recruited through social media posts. The study’s participant information was made available publicly online (https://github.com/test-trace-isolate-inte rviews/public-recruitment), and a digital copy of the participant information was distributed to participants prior to the interview. Informed consent was captured through an online booking system and confirmed at the start of each interview. Participants were offered a £50 store voucher for taking part. Demographic data was captured (gender, age, ethnicity, and whether they were working from home). No demographic quotas were set for recruitment, as the social media advertisements were placed to encourage diverse participation.

#### 2.1.2 Procedure

The interviews address the following research questions:

- RQ1: What themes emerge when the public are asked to talk about their opinions of Test, Trace and Isolate practices in the UK?
- RQ2: What factors do the public say influence people’s willingness to isolate and test?
- RQ3: What risk management strategies do people employ beyond official guidance?

Interviews were conducted with 20 members of the public, by video conference. We conducted semi-structured interviews based around a topic guide to facilitate in-depth exploration of their thoughts and gather exploratory data in an open-ended manner [21, 22]. The questions were designed to capture feed-back on the Test, Trace and Isolate processes, without requiring people to share their personal experiences, and to allow insight into behavioural practices influencing modelling decisions without requiring technical discussion of modelling with members of the public. The full schedule is available online [17].

There has been limited research on how to facilitate public engagement with epidemiological models and that which there is has noted the long time period required to develop a mutual understanding [54]. Working within a short time frame, and in a politicised media climate around modelling, we had a challenge as to how to represent TTI models to interviewees. To create the topic guide, we had multiple conversations with TTI modellers to understand priority topics, especially around assumptions made in modelling. A pilot interview was conducted using a diagrammatic stimuli based on how modellers had represented TTI process. This depiction was intended to display the TTI process as a conversation prompt; however this did not make sense to the participant, who had specific interactions with the process rather than an understanding of how those interactions fit within a more linear TTI process. We removed this stimuli and used it as part of the topic guide instead to ensure all parts of the TTI process were discussed. We also considered directly asking participants about assumptions, but shifted towards asking practically-based questions that were felt to be a more accessible way to approach modelling assumptions. Examples include delays experienced during different testing interactions, how and when they choose to test, and the barriers they see to testing. Questions were designed either to relate to assumptions made in modelling, or to provide opportunity for broader commentary on angles modellers might not have considered. We purposely did not anchor our questions in policy, but rather on experiences the participants were likely to have (e.g. attending social event).

Two researchers interviewed each participant, one following a topic guide and the other taking notes and asking any clarifying questions at the end of the interview. Each interview took approximately one hour. Following the interview, summary notes were sent to the participant, and edited and validated by the participant.

The study was conducted between 1 July 2021 and 26 July 2021. Ethical approval for this study was granted by the London School of Hygiene and Tropical Medicine (ref: 26265).

#### 2.1.3 Thematic analysis

Thematic analysis was used to analyse the interview data [9]. The analysis was data-driven, with the themes formed inductively [9]. The explicit meaning of the data was analysed, as per the realist approach, without considering the effect of socio-cultural or other factors on participants’ thoughts [9].

NVivo software was used to conduct the thematic analysis [10]. The analysis was performed according to the steps described in the guidelines for conducting thematic analysis by Braun and Clarke [9]. Two coders were involved in the analysis, with a single coder performing the initial coding of the interview summaries. The resulting codes were then refined by a second coder to facilitate validation of the coding scheme and the emergent themes. Presentation of the thematic map and the coding scheme to members of the research group was also performed to allow additional validation and crystallisation of the analysis’ results.

#### 2.1.4 Symptoms analysis

As part of the schedule, participants were asked what might make someone want to take a test for COVID-19. Participants gave a list, and these were tabulated to allow quantitative analysis. This differs to previous methods which present participants with a list to identify symptoms from [15].

### 2.2 TTI modeller input: Workshop method

#### 2.2.1 Introduction

To interpret the implications of the interview findings for assumptions made in TTI modelling, we conducted a one hour workshop with nine academic infectious disease modellers involved in Test, Trace and Isolate modelling for the COVID-19 response in the UK. This is reported in Section 5.

#### 2.2.2 Participants

Workshop participants comprised nine pandemic modellers involved in modelling testing, contact tracing and isolation policies in the UK, all of whom had been engaged with providing evidence to support policy development and were known to E.F.

#### 2.2.3 Procedure and Analysis

Results of interest from the interviews were selected (G.M, R.S, C.J, E.F) and put into a PowerPoint, with discussion points. Modellers responded to findings as they were presented and as prompted via discussion points. Notes were taken of the discussion, which was then summarised in Overleaf, a LaTeX collaboration tool (G.M, R.S, C.J, E.F). Following this initial analysis, workshop participants were offered the chance to review, comment and edit the document, through several major revisions over several months. A full list of workshop participants and the workshop materials have been made available [17].

#### 2.2.4 Consent

As part of the invitation to the workshop, it was agreed that the discussion would be analysed and written up in a report, which all participants would be invited to author. This model of collective knowledge production has previously been successfully used (e.g. [55]).

## 3 Results

This section presents the results of our thematic analysis and quantitative report on symptom identification, which were presented for further exploration with modellers as reported in Section 5.

### 3.1 Emergent themes

The emerged themes mapped to: (1) education and guidance offered publicly about SARS-CoV-2, (2) trust in the mitigating measures, (3) practical issues arising from testing and isolation uptake, (4) the mental health impact of measures for controlling SARS-CoV-2, (5) perceived risk to self and others regarding SARS-CoV-2 infection and spread, and (6) attitudes about the future and the pandemic and recommendations on controlling SARS-CoV-2 better. The relationship between themes are shown in the thematic analysis map (Figure 1).

**Figure 1:**
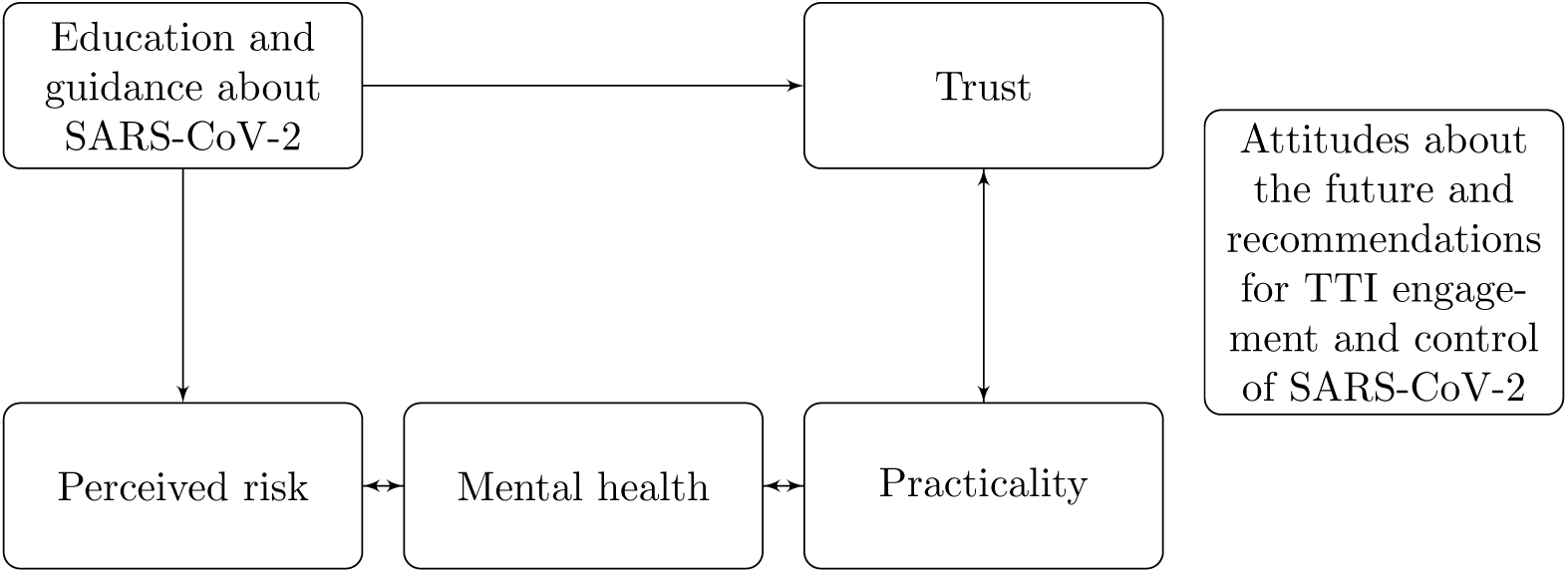
Thematic analysis map: Factors of behavioural response to pandemic and TTI guidance. The “Attitudes… ” theme was quite distinct from the other themes, and items arising as e.g. “Perceived risk” or “Mental health” were commonly not related to “Trust”

#### 3.1.1 Education and guidance about SARS-CoV-2

Participants identified what they perceived as a lack of education or official guidance about the processes of testing and isolation to influence the public’s uptake of these interventions. Reported education gaps included the current status of the pandemic, the importance and impact of SARS-CoV-2 mitigation measures, and knowledge of recommended measures. Some participants mentioned that the severity of the pandemic has not been emphasised effectively, leading to parts of the population perceiving SARS-CoV-2 as a seasonal flu. Participants reported that guidance about symptoms was unclear and that this contributed over time to symptoms being misinterpreted as seasonal colds, allergies or flu, with official guidance not effectively promoting the necessity for timely testing and isolation upon their manifestation. Participants felt that the population was unaware of asymptomatic or pre-symptomatic transmission and that this meant that the public was unmotivated to test regularly or quarantine when traced as contacts. Similarly, participants also believed that most people thought that a prior infection or double vaccination meant that you could not be infected again and so probably would not need to test or isolate as a contact.

Several participants said that some people were still unaware of or unsure about the different types of testing (PCR or LFD) and their properties, such as their effectiveness in detecting a SARS-CoV-2 infection. Lack of knowledge about how to access test kits was also perceived as a barrier to testing uptake and was attributed, again, to limited or ineffective advertising strategies. Some participants suggested that the steps that one should follow after a positive test result were unclear, including confusion about the length of isolation required, whether other members of the household should isolate as well and for how long, or how support would be provided while isolating (e.g. essential shopping delivery, childcare, work arrangements). Such confusion was thought to impede testing uptake.

#### 3.1.2 Perceived trust

Participants reported that public trust in government SARS-CoV-2 guidance, including in the TTI system, influenced uptake and adherence to TTI policies and the public’s perception of the severity of the pandemic. It was perceived that public trust in government and therefore in its competence to provide guidance had been undermined by (a) the non-adherence of prominent government officials and politicians to the rules of isolation, (b) corruption, cronyism, and profit-making of private organisations, including within the TTI system (c) confusion created by constant changing of guidance and (d) travel restrictions that seemed driven by political interests rather than the control of SARS-CoV-2. Moreover, the easing of restrictions contemporaneous with the interviews in July 2021 was perceived by participants to signify that personal adherence to the TTI processes would be *beyond what was required*, regardless of the state or trajectory of the pandemic.

Some participants commented that people’s disappointment with their direct experience of the NHS Test and Trace services when traced as contacts or when testing positive contributed to a lack of trust. A particular issue mentioned was lack of clear information regarding the isolation process, demotivating people to stay isolated, delays in NHS Test and Trace contact tracing, multiple notifications of the same contact and an impersonal attitude adopted by some call-handlers during tracing calls.

Trust in NHS Test and Trace was undermined further, according to most participants, by the perceived defective performance of the NHS COVID-19 app. The majority of participants thought that people would not quarantine if notified by the NHS COVID-19 app due to its perceived limited specificity. Mistrust in the NHS COVID-19 app notifications arose due to people receiving multiple “pings” requesting the recipient to quarantine or, conversely, not receiving “pings” when they knew they had been in close contact with people who had tested positive. Other technical issues were mentioned as reducing trust, particularly methods for avoiding or removing isolation notifications, such as the ability to delete check-ins, re-install the app or to simply avoid using the app. A lack of consistency with Test and Trace calls concerning the length of the isolation period was also viewed as reducing trust in the tracing process and subsequently testing and isolation. Privacy concerns about personal data handling due to the perception that data was not being anonymised were also perceived by participants to prevent app usage.

Participants noted that contact tracing also occurred informally without the involvement of NHS Test and Trace, such as via WhatsApp group messaging with people they knew, though they had mixed views on whether this informal tracing was more or less trusted than official channels. Several participants viewed NHS Test and Trace and the NHS COVID-19 app as conceptually appropriate, but felt they could be more effective in controlling SARS-CoV-2 if applied in a more systematic, consistent and organised manner, and if compliance checks were in place. Some participants added that these services did their best in reacting quickly, considering that the pandemic was an unprecedented situation. Specific recommendations for improvement mentioned by participants are captured in Section 3.1.6.

Concerns about trust were also mentioned with regard to testing technologies. Almost all participants perceived PCR tests as more accurate than LFD tests, and believed that the majority of people would isolate following a positive PCR test result. In contrast, participants thought that people would not entirely trust a positive LFD test result and would want to validate the result via a PCR. The mistrust in LFD tests was not symmetrical: most participants suggested that a negative LFD test result would mean that the person had not been infected with SARS-CoV-2 and would not continue to isolate when traced as a contact of a confirmed case. Negative publicity around inadequate testing capacity and the reporting of SARS-CoV-2 cases, particularly on television news, was thought by a few participants to undermine trust in the testing process and test results, in turn reducing willingness to isolate. Reporting of the LFD test results was also linked by a few participants to privacy concerns about data handling, suggesting that this would make people less willing to share their test results.

‘Surge testing’ interventions, in which whole communities were asked to test in response to variants of concern in the first half of 2021, invoked mixed reactions. Some participants raised privacy concerns in relation to door-to-door testing and thought that some groups with previous negative experiences of governmental control, such as refugees, might feel intimidated. However, most participants held positive attitudes towards surge testing, acknowledging its usefulness for identifying cases and new variants. Participants also mentioned the social benefits of surge testing, as it feels more like a local community effort to reduce spread.

#### 3.1.3 Practicalities

Participants collectively acknowledged that attitudes to testing and isolation strongly linked to practical issues. Almost all participants noted that missing work and the related financial implications would be a common concern for those who were required to isolate or quarantine. More specifically, participants mentioned that a lot of people would fear the possibility of losing their job if they isolated at home, especially if they were required to do this multiple times or for long periods. Several participants also commented that some employers were pressurising people to go back to work, even if they tested positive, and reported employers suggesting ways to circumvent the isolation policy after a positive test result, such as deleting the NHS app in order to erase the isolation notification. Some of the participants in the study were healthcare workers, and they mentioned pressure to stay at work, even if traced as contacts during their shift, due to the increased need for medical personnel during the pandemic. In addition to increasing pressure on co-workers, participants mentioned that people would worry about losing their salary, especially if they were on hourly contracts, or paid less while isolating. For these reasons, participants thought that people would be less inclined to test when relevant, to reduce the chance of being told to quarantine. A few participants, however, held positive views about working from home, as the nature of their work allowed this; these participants also had a more positive attitude towards testing and isolation.

Participants reported particular practical challenges for parents or caregivers, which they thought could influence their uptake and adherence to TTI policies. Adherence to isolation was thought to be negatively influenced by an incompatibility of working while providing childcare in self-isolation at home or with an inability to look after elderly, disabled or vulnerable individuals. The ability to isolate parts of a household within a home in any of the contexts above was thought to ease the impact of isolation, depending on people’s living conditions, such as the house layout and size of household. Those with more physical space would find it easier to partition the household into COVID-positive and COVID-negative subsets for within-household care.

Child testing was additionally considered by participants to have practical issues, in terms of making time in daily routines to do the tests and ensuring older children were doing them regularly as instructed for school. Another challenge noted by parents was the logistics associated with taking a PCR test in order to get young children back to nurseries. The negative impact of negotiating the logistics of testing was thought to be exacerbated when parents perceived that their child’s symptoms had other causes (e.g. fever caused by teething was mentioned). Additional issues included the difficulty of performing swabbing on children, and the trauma this could cause. A few participants viewed testing children to be unethical and coercive.

Convenience and availability of testing was perceived to affect uptake among adults as well as children. Discomfort with swabbing was thought to reduce willingness to test. Some participants also expressed concerns about whether they were performing the test correctly (on themselves or on children). Home test kits (both PCR or LFD) were considered collectively by participants to enhance testing uptake, particularly when there was convenient access to free LFDs. Some participants were unsure (or mentioned others would be unsure) of where to get LFDs or the circumstances under which they would be entitled to a PCR test. Convenience also seemed to be linked to whether people were required to test regularly for work purposes (and were given LFDs) and whether they knew the distinction between LFD and PCR tests. Convenience and being free was thought to prompt people to test more often, whether it was a work requirement or not. Most participants suggested that they would opt for an LFD test if they had SARS-CoV-2 related symptoms or if they were traced as contacts due to it being more easily accessible, and then taking a PCR in a test centre if wanting to verify a positive LFD test result. Access to PCR test centres was considered to depend on geographical region, affecting both testing uptake and the speed of response to a requirement to test. The time taken to get PCR results was also a concern. Whether or not people reported an LFD test result depended on the ease of reporting it, though if the test result was positive they would be more likely to report it. Some participants found the reporting process straightforward while others described it as time-consuming or had technical challenges.

Not wishing to impact future plans, such as travel arrangements, was thought by some participants to reduce willingness to test, as it brought with it the possibility of having to isolate and cancel current arrangements. Conversely, travelling internationally was thought to increase people’s willingness to test, as it was necessary to comply with many countries’ border controls. Similarly, the requirement of a negative test result to access some venues or events was seen to increase testing uptake. Other practical issues were language barriers, which hindered people’s understanding of Test and Trace phone calls and government guidance. Finally, the perceived effectiveness of contact tracing strategies in reaching people was identified by several participants as a factor underpinning limited testing and isolation uptake. Limited access to the NHS app due to people not owning or knowing how to use a smartphone, such as some elderly people, and lack contact tracing process guidance for children were perceived as contributing to this. Delays in the TTI system, such as waiting for the test result or waiting for the complete list of contacts to be contacted by NHS Test and Trace, was perceived by some participants to affect timely testing and isolation.

#### 3.1.4 Mental health impact

Mental health was identified by participants as an important factor influencing people’s attitudes and adherence to TTI guidance. Participants collectively highlighted that people were fed up with the pandemic, and particularly with isolating and missing out on social interaction. This was considered to reduce the willingness to isolate. It was also considered to reduce willingness to test, as some people would be hesitant to identify a potential infection in themselves. The impact of isolation on mental health was discussed in detail by several participants, who suggested that if people live in a small apartment without a garden and had experienced challenges isolating multiple times during lockdowns, they would feel challenged mentally to do so again if traced as contacts. People’s perceived need to isolate would also be reduced by testing negative via an LFD or after having achieved a level of immunity through vaccination or prior SARS-CoV-2 infection. More broadly, people found isolation more difficult mentally when they did not believe that there was a strong likelihood they had really been infected. Some participants mentioned that it would be even more challenging for people with pre-existing mental health disorders to keep isolating, if previous isolation periods had exacerbated symptoms. A few participants noted that apart from isolation and the repercussions associated with it, testing could also be a stressful procedure for some people, eliciting anxiety about whether they would test positive and isolate while worrying about their physical well-being. People who had never tested before expressed anxiety about performing the testing procedure itself.

Testing with LFDs in particular also had a positive role to play in reducing anxiety, as it allowed people to check their own infection status when traced as contacts, or if they had SARS-CoV-2 related symptoms. People valued LFD testing when meeting vulnerable people, as it allowed them to be more confident they were not going to spread infection, with subsequent positive effects for mental well-being. The same was thought by participants to apply to those having had a bad experience of prior SARS-CoV-2 infection. Most participants mentioned that people who were themselves severely ill with COVID-19, or knew people who had been, would be more inclined to test and isolate, to reduce their anxiety of themselves or others experiencing the negative impacts of the illness.

#### 3.1.5 Perceived risk

Perceived risk to self and others was seen by participants as a factor in determining testing and isolation uptake. More specifically, the extent to which people thought themselves to be immune was thought to affect their response when traced as contacts or when presenting SARS-CoV-2 related symptoms. Those believing themselves to be at low or no risk of infection would be less likely to test or isolate. Immunity was considered to be achieved either through vaccination or prior SARS-CoV-2 infection. The majority of participants mentioned that whether people would feel protected and subsequently test and isolate would depend on their personal mindset and relevant education, although vaccination was perceived as more protective than immunity provided via prior infection. Some participants said this was due to the unavailability of data linked to the possibility of re-infection.

Participants felt that an individual’s mindset also affected their perceived risk of being infected. For example, most participants suggested that people who did not believe in the existence or severity of SARS-CoV-2 would not be inclined to test and isolate in any scenario, nor vaccinate. Similarly, religious and cultural reasons were thought by a few participants to shape attitudes towards testing and isolation and subsequent uptake. Being younger was also perceived to result in reduced likelihood of testing and isolation due to the belief that young people would be less vulnerable than older individuals. In contrast, a few younger participants suggested that elderly people were showcasing risky behaviour after vaccination, wanting to travel before everyone would be vaccinated.

Testing interacted with personal risk assessments. A negative LFD test result was suggested by several participants to reduce adherence to isolation when people were traced as contacts or when they had SARS-CoV-2 related symptoms. Almost all participants said a negative PCR test meant a person did not have COVID-19 and thus would not quarantine. Correspondingly, not having symptoms, or not having severe symptoms, even when someone had tested positive for SARS-CoV-2, was considered by some participants to reduce testing and isolation uptake. The risk of being infected with SARS-CoV-2 while queuing in a test centre was thought by a few participants to demotivate people to test when a PCR test was required.

The perceived duty to protect others was also seen by participants to influence testing and isolation uptake. All participants mentioned that they would test before visiting vulnerable people and family members, especially those who were elderly or young children. Correspondingly, those who were not living with vulnerable people or meeting non-vulnerable peers were less likely to engage in testing and isolation, resulting in riskier behaviours. Finally, the feeling of personal responsibility to protect the community in general was identified by some participants as a reason for people to test as per official guidance, irrespective of whether they were meeting vulnerable people. The community impact was also discussed by several participants as a motivating factor for testing and isolation uptake in scenarios like surge testing and where non-adherence to guidance might be negatively perceived by others.

Perceived risk was seen to change according to the phase of the pandemic. Most participants suggested that during periods of fewer cases, people would be more relaxed and, thus, less inclined to test and isolate. Similarly, some participants mentioned that people were less willing to test and isolate after the first wave as morbidity and hospitalisation numbers were thought to be reduced and the social impact of spreading COVID-19 was not as severe as previously. Summertime and good weather were additionally perceived by participants to introduce riskier behaviour due to increased socialising and less motivation to test and isolate as this could lead to missing out if one tested positive for SARS-CoV-2. The majority of participants also explained that the ongoing increase in vaccination numbers offered people a sense of safety, reducing their perception of the necessity of testing and isolating if they had relevant symptoms or were traced as contacts. The spread of new variants was a factor that for some participants would increase testing and isolation uptake, as under these circumstances the effectiveness of vaccination was less clear, and there was potentially a greater threat to people’s health. Finally, the easing of restrictions in July 2021 was thought by several participants to suggest that testing and isolation would not be needed as everything was going back to normal and the pandemic had ended, reducing people’s perception of the severity of the situation.

#### 3.1.6 Attitudes about the future and participant recommendations

Participants’ views regarding both the evolution of the pandemic and the viability of the TTI guidance were mixed. Some participants believed that most people were optimistic and less scared than previously, thinking that the pandemic would soon be over and restrictions would progressively ease. Others suggested that the pandemic would not end in the near future and they would continue to maintain a cautious lifestyle, perceiving some restrictions as necessary. Some participants also mentioned that they were happy with some restrictions remaining, as it made them feel more secure, enabling socialising and preventing future lockdowns. For these people, the easing of restrictions was seen as irrational. Regardless of the number of SARS-CoV-2 cases reported, practical issues such as the repercussions of testing and isolation on work and the negative impact of the restrictions on the economy were thought to thought to make people more likely to believe that restrictions should be lifted and future lockdowns avoided. Interestingly, while most participants acknowledged that a large proportion of people no longer wanted restrictions, as they were fatigued with the situation and had been vaccinated, they suggested that they would continue to lead a lower risk lifestyle and abide by measures for the control of SARS-CoV-2 like mask wearing, hand sanitising, testing and isolating.

As discussed previously, the quality of the government’s response to controlling SARS-CoV-2 spread also influenced people’s attitudes towards the pandemic and to mitigation measures, as did positive views about surge testing, and are included in this theme.

Several participants mentioned the need to learn lessons from this pandemic. One group of participants mentioned the government’s responsibility to understand how to improve its response to future pandemics. Others commented on the opportunity offered to learn from this pandemic and the need to realise personal responsibility in tackling this kind of situation, where people were members of a community, whose actions affected each other.

Participants also provided recommendations on how to improve policies for controlling SARS-CoV-2 including increasing engagement with the TTI system and adherence to guidance. These included: support from both a financial and mental health perspective during the isolation period; follow-up advice on the length of the isolation and next steps; incentives such as food vouchers, paid self-isolation support, or paid leave. Raising awareness about the severity of the pandemic and the importance of personal responsibility through education about the virus, and advertising testing availability and the steps required following a positive test or when traced as a contact were additionally thought to promote people’s motivation to test and isolate. This also applied to promoting vaccination uptake. Scientists could play an increased role in communicating their knowledge about the virus and improving trust in the measures recommended for controlling SARS-CoV-2. It was also suggested that social media campaigns and influencers could raise awareness about personal responsibility to protect others and improve adherence to testing and isolation.

Ensuring testing availability was thought to motivate uptake. Developing new means of testing that were less traumatising for children was also suggested. On-site testing in schools was seen by parents as positive, as it avoided the difficulties of testing (and remembering to test) children at home. Some participants felt that making testing mandatory would increase compliance. Similarly, workplaces complying to testing and isolation guidance was thought to improve uptake. Some participants thought shorter isolation periods could be traded off against greater testing as a way of increasing isolation uptake. Correspondingly, shorter isolation periods for those who had been vaccinated was also recommended.

Other measures to control the spread of SARS-CoV-2 that participants found acceptable were mask wearing, hand washing, social distancing, not sharing drinks, eating healthily, taking one’s temperature upon entering premises, maintaining bubbles and meeting in well ventilated places. Testing before big events and testing visitors in care facilities was also perceived by some participants to help protect vulnerable individuals. Limiting travel to other countries was recommended by a few participants to help prevent the spread of new variants. Finally, new treatments for COVID-19 and promotion of vaccination were considered solutions to COVID-19 regardless of its prevalence.

### 3.2 Symptom identification

Participants were asked “what symptoms might make someone want to test for Covid (SARS-CoV-2)”. This was not incorporated into thematic analysis, being quantitative, and is reported separately in this subsection.

10/20 participants mentioned all three of the major symptoms (cough, fever or high temperature, loss of taste or smell). Except for three participants, who mentioned precisely the three main symptoms with no additional symptoms, no two participants listed the same symptoms as one-another. The other symptoms mentioned by participants in various combinations were headache, sore throat, upset stomach, flu-like symptoms, feeling generally unwell, fatigue or tiredness, back pain, runny nose, skin rash, aches and pains, “lots of other symptoms”, and a head rush. Two participants said that having a fever would make people *not* want to take a test, since they would think the symptom more likely to be caused by a heavy cold or flu.

## 4 Discussion

The interviews identified a range of themes emerging from reports of the public’s experiences and perceptions of Test, Trace and Isolate strategies for COVID-19 control in England over 2020 to July 2021. In this section, we first reflect on the similarities and differences in the themes our study has identified compared to others, develop our workshop discussions about what these findings mean for how TTI is modelled for informing policy, review the strengths and limitations of our approach and suggest the implications for pandemic transmission and inetrvention response modelling in future.

### 4.1 Comparison to previous literature about public engagement with TTI and other COVID-19 control policies

**Lack of guidance** or confusion with government policies related to COVID-19, including but not limited to those relating to TTI, have been reported in other qualitative studies and behavioural surveys. Previous studies of COVID-19 pandemic control measures including social distancing and isolation during March-April 2020, the time of the first ‘lockdown’ in the UK, found similar themes despite the earlier stage of the pandemic [2] and the setting prior to the establishment of NHS Test and Trace in May 2020. Themes emerging from focus groups at this time included loss (social, financial, mental, self-worth); criticism of government communication; adherence, including “High levels of self-adherence but observations of non-adherence in others” which was also identified in our study; and uncertainty. A follow-up study focusing on non-adherence to control policies was conducted in September-November 2020, still nine months prior to ours, and identified some additional themes which again were emerged in our study, including lack of trust in government (absent from the earlier study) and perceived inconsistencies, and fatigue with and difficulties in following changing guidance[3]. Specifically with regard to TTI policy, the CORSAIR study also identified confusion as to guidance, including in identification of symptoms meant to prompt isolation and testing, lack of clarity as to testing policy (e.g. when to use LFD tests versus PCR tests and interpretation of their results) [6]. Our study found similar confusion but also that people’s choice of test was based primarily on convenience, accessibility, trust in tests, and whether they thought they might have COVID-19.

**Lack of trust** has also been found in other UK studies, including in interviews conducted amongst members of the public in Sheffield, England from October 2020 -February 2021, with people wanting further evidence as to their risk of infection as close contacts asked to quarantine [5]. At the time of our interviews in July 2021 we did not find people wanting further evidence but rather making their own risk assessments. Along with another study from the UK in from November/-December 2020, we did find that individuals tended to be more inclined to test and isolate when experiencing symptoms [38] and people felt encouraged to test when they thought they had COVID-19. A study reporting that “participants did not accept the result of the LFD as conclusive evidence that they did not have the virus” [4] echoes the lack of trust in LFDs expressed by participants in our study

Our findings align with others identifying **practical constraints** to adhering to TTI policies, many of which disproportionately affected those in less social and economically secure and with caring responsibilities. The COVID-19 Social Study found heterogeneity in uptake and adherence to isolation policies during Winter 2020-2021 [39]. Another study found that risk factors for non-adherence to self-isolation rules include low income backgrounds or lack of paid time off [59]. This was also a topic mentioned by participants of our study. The concordant inequalities in financial challenges and self-reported lack of adherence, including in the UK, has also been well documented and used to inform policy (see e.g. [44]).

Our study found many concerns expressed about **mental health impacts** of TTI policies. This is consistent with what others have found. Across the world, the pandemic has had a negative impact on mental health, and has been linked to increasing cases and severity of depression, anxiety, and traumatic stress (e.g. [58]). See Boden et al. [57] for further references, including the impact on vulnerable populations.

Self-isolation during the pandemic has been seen as purposeful or forced, relating TTI to mental health [8]. The main mental health topics found in our study have been highlighted by others: Cultural differences[31], differences in support networks[31], family support commitments[34], and differences in financial impact were found to be important in our study. The UK COVID-19 Social Study found mental health issues, such as depressive symptoms, to be “significantly higher among people experiencing abuse or low social support, individuals with low socioeconomic position, and those with preexisting mental and physical health conditions” [20]. They suggest that mental health and socioeconomic interventions should be targeted towards people with these risk factors. This matches well with our findings of public perception of those finding it most difficult to isolate, perhaps with the addition of those with families as a priority group for mental health targeting.

Isherwood et al. [40] found, in Wales, “Self-isolation is particularly challenging for younger people, women and those with precarious incomes”. As with our study, they found a barrier to self-isolation was wanting to see family and friends. Being a parent/carer was not captured as demographic data, but may be correlated with demographics found to be more likely to find self-isolation challenging and to report mental-health issues (younger, women, precarious income).

Various scales exist to examine **risk perception** associated with COVID-19, including CORAS which was validated in the UK [60]. CORAS distinguishes between likelihood and severity, and evaluates an initial nine item list to be self-rated, such as “feel vulnerable”, “concern about getting infected” and “gut feeling of own likelihood of infection”. Their items are broad, but cover topics mentioned by participants in our study. In a spring 2020 survey, Dryhurst et al. [19] found that risk perception of SARS-CoV-2 was higher in the UK than in other countries studied. Together with the different policies adopted through the pandemic, this could make the results about changing risk attitudes and barriers to testing/isolation found in our July 2021 study less applicable to other contexts.

Part of the risk assessment described by participants in our study was about protecting older or vulnerable relatives, though this was strongest with respect to specific personal relationships. This was also reported by elder adults in Switzerland in April-May 2020 [27], though both studies found some intergenerational tensions resulting from COVID-19’s strong age-differentiated risk of severe disease; older people criticised young people in both settings, though we also found young people criticising older people for demanding to go on holiday before vaccinations were made available to younger people.

Our results found that some parents found testing on children to be unethical and coercive. This is interesting because of the perception (and actuality) that children are at very low risk of disease even if infected. This raises a question about the motivations for parents conducting non-mandatory testing in preschool children at all, which could be about desire to conform to guidance, selfishness (i.e. am I going to get ill) or a desire not to spread the illness. For children, the low risk and impact reduces the value of testing, so people were implicitly (at least) valuing the benefit to others (including themselves) against the negative impact of performing the test on children. For children, the risk and impact of SARS-CoV-2 is low with currently known variants. We might expect attitudes to child testing to change if there was a significant disease risk for children either from a new variant or the next pandemic.

In our study, participants felt that vaccination would have a different effect on risk profile and behaviours to being previously infected with COVID-19. However, from a risk-basis, prior infection has been shown to provide protection against future infection with SARS-CoV-2, to the extent that it has been advised vaccinations can be prioritised to other groups [33]. Participants felt that the severity of experiences of those having the infection was more important than prior infection providing protection, in terms of people’s risk management and impact assessment.

Our findings regarding **symptom identification** align with the findings of other studies in the UK. Through 37 national surveys, Smith et al. [15] found that 51% of participants were able to identify the main symptoms of COVID-19. They “identify the most common symptoms of COVID-19, with multiple response options allowed (up to four initially, up to five from 25 May 2020, wave 18). We coded participants as having identified symptoms of COVID-19 if they selected cough, high temperature or fever, and, from 18 May 2020 (wave 17), either loss of sense of smell or loss of sense of taste. In government guidance these symptoms are actively promoted to members of the UK public as the “main” symptoms of COVID-19” and are intended to prompt symptomatic testing via PCR. Our finding that 50% of participants listed all three of the major symptoms matches closely to this survey synthesis, showing robustness across different surveys and specific methodologies and over time. Beliefs about symptoms, and the heterogeneity of these beliefs, has a direct impact on individuals’ testing and isolation practices, as it drives whether they think they may be infected with SARS-CoV-2.

The mixed views found in this study about what symptoms would make someone want to take a COVID-19 test suggests either that symptom guidance is not being well communicated, and/or that personal experience, knowledge or risk-profile are playing an influential role in whether someone might take a test when presenting particular symptoms.

Several studies (e.g. [61, 62, 63]) have considered the effects of changing the symptoms that prompt symptomatic testing, e.g. by adding additional symptoms onto the symptomatic testing criteria. Given the already high levels of confusion, changing the policy might not improve case detection in practice without attention to also addressing other behavioural and structural reasons for non-uptake of symptomatic testing. Several participants suggested that going to test centres was undesirable, and that the ease of access to LFD tests made them a desirable choice. Symptomatic testing then may be improved by endorsing the use of LFDs or pre-delivering PCR tests to households for use when they develop symptoms, as this would lower the barrier to symptomatic testing, and would remove the delivery delay from home test kits.

**Perception and usage of different test types** has emerged from a few studies in the UK. A negative LFD test result was more likely to be trusted than a positive LFD test result, despite their high specificity and a greater epidemiological concern about low test sensitivity to early infection [51]. It is possible the trust in LFD tests, at least in part, is due to how test results are managed: a positive LFD has meant that people then need to get a PCR test, but a negative LFD means you can visit a care home. The logical conclusion of this policy/management is that a positive result needs confirmation, but a negative result can be trusted. The perception could also be due to concerns given media attention in Spring 2021 over false positive LFD tests [68].

Participants reported low trust in the accuracy of LFDs, and at the same time thought people would not quarantine if they had a negative LFD test result. This could either be because they believe their risk to be lower (even not trusting LFDs entirely), or because people do not want to isolate and are performing “mental gymnastics” in order to avoid quarantine. The “false sense of security” given by false negatives has been a highlighted as a concern by Public Health officials [42].

LFD trust may be low due to various factors, such as media influence, that PCRs were deployed first, that official bodies often require PCR rather than LFD results, and that LFDs are available for free in the UK.

Participants in our study described a range of what could be considered **“Community driven” prevention strategies**. Many of the additional SARS-CoV-2 prevention measures taken by the public, such as using LFDs before meeting vulnerable relatives, represent the outcome of the public’s understanding of COVID-19. their risk perceptions and their assessment of the coasts and benefits of the measure compared to their risk perception. As well as providing examples of measures which could be seen as feasible and acceptable to at least some members of the public, these measures give useful information about the trade-offs that the public is willing to make and highlight what they value. These insights are valuable to understanding what interventions are likely to be effective in practice, and in suggesting approaches that might not have been apparent to modellers, or to public health researchers and policy-makers considering policy options.The usefulness of community-driven prevention strategies has been strongly highlighted in experience with HIV for example, for instance with regards to ‘serosorting’ among men who have sex with men to reduce the likelihood of transmission. This was not originally a ‘top-down’ public health initiated approach to reducing risk. It may be possible to consider similar approaches in future pandemics.

Lucas et al. [16] suggest that the duration of self-isolation should be weighed against the reduced self-reporting rate associated with longer mandated duration of self-isolation. That one of our participants described being told to quarantine, isolate and quarantine for six consecutive weeks, subsequently choosing not to adhere to an isolation instruction to attend an important family event, supports this suggestion. Again, working with the public to understand what is reasonable policy to ask people to comply with may result in improved adherence. To improve the effectiveness of this action, it may be worthwhile to explore interventions to explain viral load and communicate about effective test timing.

## 5 Reflections on implications for modelling

This section presents some specific findings of consequence to modelling decisions, and ways in which the qualitative data gathered in this study could be of relevance. Many findings have implications for how modelling results should be interpreted in policy-making, and highlight the need to place transmission modelling within a broader framework to consider benefits and harms of different policies, for different population groups; here we highlight particularly the implications of findings for choices made in the process of modelling TTI policies on SARS CoV-2 transmission. The insights in this section were gathered through a workshop with pandemic modellers, as outlined in Section 2 2.2.

Some of the findings have implications for the **structure and types of models** used to investigate the effectiveness of TTI policies, while others inform likely **parameter ranges** or distributions, the choice of which has been shown to be one of the main factors in estimation of TTI impact[70]. In some cases, findings help to inform **likely trade-offs** that models could be employed to investigate, as was done in a previous analysis of the potential effects of mandated isolation depending on how this affected testing behaviour [16].

The interview findings have numerous implications for how individual behaviour is incorporated into models, from **how behaviours at different stages of the process are correlated** with each other to **how behaviour might change over time** or as the result of other experiences, and to the extent to which individuals’ behaviours are associated with those with whom they share a household, or make regular contact, leading to **clustered behaviour** across the population. Differences in the way these behaviours are represented both spatially and demographically could have important epidemiological consequences.

Finally, our interview findings **highlight research questions** that follow from how people are actually engaging with TTI in practice. The strategies that they adopt result from individual attempts to balance the costs and benefits of engagement and highlight key situations in which members of the public require guidance. We can therefore learn from these strategies and consider how further modelling research could be used to support them.

### 5.1 Informing TTI engagement estimates chosen in model parameterisation

#### Symptom identification, testing and isolation uptake

Approximately 50% of participants did not mention all of the main three symptoms, and there were 18 different answers from 20 people about what symptoms might make someone want to take a SARS-CoV-2 test. This suggests that the symptoms communication since May 2020 [15] has not been particularly effective, and that many people with COVID-19 symptoms in July 2021 would not recognise them as such. Children have different main symptoms to adults (headache, rhinorrhoea and fatigue) [69], and it is possible this is also a point of confusion.

TTI intervention effectiveness is sensitive to the proportion of symptomatic cases who go on to isolate and test [7, 70, 16]. These actions depend upon recognition of which symptoms are intended to prompt this behaviour and are a key parameter in modelling of symptomatic testing strategies. As well as suggesting the need for improved public health communication, a lack of clarity around which symptoms initiate testing could suggest that a lower range of assumed values for uptake of testing or of proportion of cases detected could be warranted. On the other hand, inaccurate application of the symptomatic testing criteria in practice could suggest that estimates of assumed test specificity and therefore of projected demand for testing could be inaccurate. Both of these outcomes could be important for informing the likely impacts of different TTI policy choices in practice.

The lack of understanding regarding asymptomatic transmission could imply that adherence is poor while an individual is asymptomatic, but better while they are symptomatic, which could be incorporated in and explored within a TTI model that stratified cases by these criteria, or potentially linked to symptom onset time where this is assigned for modelled cases, as discussed by Quilty et al. [65].

#### Variations in testing accuracy and behaviour

Studies of test sensitivity suggest that test performance depends in part on the training of the person performing the test [51]. This can be incorporated into testing models by incorporating a ‘swabbing error rate’ in addition to other factors having a bearing on test sensitivity, most notably viral load at time of testing. However, our interview results also suggest that this error rate might vary by age of the individual being tested given difficulties with testing young children, the effects of which could be explored in modelling by varying the swab error rate. This could be important for exploring varied effects of testing within different age specific settings such as schools or care homes. Variation by age in test booking conditional on symptoms could also be explored further, given the mentioned difficulties and perceived unpleasantness.

#### Test result interpretation and subsequent actions

A major question for UK TTI policy has been the effective use of lateral flow testing, which instead of detecting viral RNA, as a PCR test does, identifies viral antigen. PCR testing is more sensitive, identifying individuals with lower levels of virus both at the beginning and end of their infection, but requires laboratory analysis, increasing costs and time to return results, and may detect virus in individuals who are no longer infectious to others. LFD tests are less sensitive to identifying individuals who have lower viral load, which includes both those at the very beginning of infection, prior to becoming more infectious, and at the end of their infection, when they likely are not infectious. However, studies such as Pickering et al. [66] and Lee et al. [67] have found high viral loads closely correlated with LFD positivity, the presence of live (infectious) virus, and onwards transmission, indicating that LFDs may be effective in reducing transmission from these individuals. Modelling has considered how the tests might most usefully be deployed, reflecting the properties of each, in order to reduce population transmission whilst reducing isolation costs among people not infected. However, there has been relatively little data available to understand how people interpret the tests in practice.

Neither a PCR nor an LFD test can definitively rule out infection at a single time point, either because the swab may not have been performed correctly, or an individual might be at the very beginning or end of their infection, when their viral load is too low to be detectable on either test, though this problem is more significant for LFD for which viral load must be higher. Our interviews suggest that messaging about the insufficiency of a single test to indicate non-infected status has not been sufficient. Almost all participants said that people would not quarantine as a contact after having a negative lateral flow test result (during a period when this was still required, regardless of vaccination status). Behaviours such as the lack of reporting negative LFD results described in this study can be seen in Public Health data [6], and combined with the above non-quarantining behaviour these cases would be difficult to monitor. Additionally, since a number of participants mentioned that they would take a home LFD test before a PCR if they developed symptoms, this raises additional concerns around case identification if false negative LFD results are then being used as justification to not pursue further testing or engage with TTI recommendations.

This finding should be incorporated and explored in TTI modelling, to assess the extent to which it could contribute to onwards transmission. It also suggests that policies that require individuals to both test and isolate (or just to isolate but in a context in which tests are made available), are unlikely to be adhered to, and that alternatives, such as daily testing amongst those at higher risk of infection, might be preferable. This point is important, because previously modelling found that the most theoretically effective TTI strategy would be to ask household contacts to both test and isolate [36], but only assuming complete adherence to the policy. Our findings support the notion that this is not a realistic assumption and that alternatives such as daily testing of household contacts, along with non-household contacts, could be more acceptable amongst the population and potentially more effective overall [37, 65].

### 5.2 Incorporating clustering and heterogeneities across the population

Cultural and religious differences, together with socio-economic and educational differences, were mentioned by participants as having an impact on willingness to test and isolate. The need for support to isolate and the heterogeneous impacts of TTI requirements to isolate on job performance and security were both mentioned. Participants discussed lack of education as being a barrier to testing and isolating, and clustering on household education level would be possible. Participants also described how large families may be more likely to be asked to isolate (participants used the word isolate but perhaps meant quarantine) repeatedly, and be less able to adhere to that guidance.

People sharing socio-demographic characteristics within a geographical area are more likely to be clustered together on a dynamic network of social contacts via household, family, workplace and friendship relationships. When these socioeconomic and cultural characteristics are associated with uptake and the ability and/or willingness to engage with contact tracing and isolation, this means that engagement with TTI is not randomly distributed across the population, but is clustered together. People who struggle to adhere are likely to be making contact with others in a similar position, and vice versa. If the factors that make engagement with TTI more challenging for some groups also hold for other interventions and for vaccination, there is the potential for ‘pockets’ of increased risk of transmission within the population to develop if TTI interventions are not tailored to the needs of these populations. This clustering would be further exacerbated if also associated with vaccine uptake, and has implications for expected transmission dynamics and interpretation of surveillance data.

Exploring the effects of clustered TTI engagement on epidemic dynamics can be done explicitly in a network or individual-based model. Alternatively, to explore how TTI effectiveness might vary among sub-populations, a simpler model could still be informative drawing on different parameter value ranges to produce sub-populations with heterogeneity in TTI engagement, for instance, likelihood of symptom recognition and testing or probability of effective isolation. This analysis, while not reflecting dynamics between population sub-groups, could in some cases be informative as to the differential impact that policies might have across different groups in the population.

### 5.3 Time-based behavioural changes

Compared with earlier studies conducted in 2020 [15], our 2021 study reports more long-term “covid fatigue” and a weariness of constant adherence to guidance.

The relationship between time spent under social distancing restrictions and testing, tracing and isolation protocol and willingness/capability to adhere to guidance has been contested over the period of the pandemic. Our findings do suggest a weariness with complying to policies and highlight the costs associated with them and with the crisis as a whole, reflecting concerns about poor mental health and isolation. There are a number of adherence ‘decay’ function options that could be incorporated in a model to assess how a waning of adherence to TTI and other policies might affect transmission and disease outcomes over a longer time frame, and heterogeneity in the extent of waning could also be considered. The translation of interview findings to the most appropriate quantitative expression of this phenomenon would still be challenging. Given the significant length of a pandemic, however, a policy that provides minor gains in reducing transmission may perform much worse in the long term, if it marginally increases the rate of adherence ‘decay’. As such, these considerations could play a role in sensitivity analyses of models.

In addition to a ‘decay’ in adherence willingness over time due to fatigue, our findings show that adherence is strongly linked to changing risk perception over time – both to the individual and to the population as a whole. This could be incorporated into modelling in a number of ways, such as by attempting to quantify a negative correlation between vaccination coverage and adherent behaviours, or by considering the impact of changes in daily case numbers. This could be modelled at an individual and a population level, with vaccinated individuals behaving differently to non-vaccinated individuals, but also by considering temporal changes in overall adherence of the population based on situational factors. Public awareness of the waning effect of vaccination and new variants would mean that ideally models would be able to also consider changes in the perceived personal infection risk, and subsequent behaviour, depending on temporal factors such as time since vaccination.

Depending on the modelling question, the relationship between behaviour and previous personal experiences, such as severe illness among loved ones, could also be included and explored in individual-based models. The effects of previous disease experience on behaviour could also be translated into assumptions about average adherence in hard-hit sub-populations versus those whose disease burden has been relatively light. However, other constraints on adherence behaviour, such as insecure employment, would also need to be considered. This effect could be analogous to models that consider a link between epidemic trajectory/severity and dynamically changing behaviour in the population in response, though these have been challenging to parameterise [25].

### 5.4 Identifying research priorities to support personal risk management strategies

Most participants wanted to conform with guidance, but only when it matched what they felt was sensible. Additional strategies not included in guidance but described by participants were (a) partitioning the household into two when someone has tested positive for SARS-CoV-2, particularly in families with children (b) testing before visiting vulnerable people and (c) continuing to comply to previous guidance after the rules have been relaxed. Given that these behaviours reflect strategies that the public has shown to be willing and able to employ, it makes sense to investigate their likely efficacy and how they could be improved upon. These behaviours could be modelled, using for example household density as a proxy for ability to partition households. The “partitioning household behaviour” would be relevant for out-of-household isolation strategies, but would probably be too small of an effect to include in most models.

#### Pre-contact testing

Many participants reported taking tests before visiting vulnerable people, yet there is not specific advice to do this nor about how to do this optimally to reduce risk of transmission (e.g. timing of test, testing technologies). The adoption of the approach into policy or as official advice would ideally be further supported by empirical evidence that reflects effectiveness of testing in practice, for instance, or ’risk compensation’ behaviours. However, that the public is using tests in this way suggests a need for guidance and public health tools in supporting people to see more clinically vulnerable friends and family and helps to prioritise modelling research questions.

#### Informal tracing

Interview findings also suggested that some of the population was informally conducting contact tracing amongst themselves, for instance via private WhatsApp groups, which may be considerably faster than through official channels, a key factor determining the success of TTI. The inclusion of informal behaviour, or behaviours not directly related to policy, is important for predicting the impact of policies. It could be difficult to quantify and characterise additional informal tracing within a model and our interviews suggest that its effects on behaviour and transmission could be heterogeneous, but awareness of its existence could aid in interpreting observed trends.

#### Supporting personal cost-benefit trade-offs

From this and other studies, it is clear that intervention design decisions should assume that adherence will not necessarily be 100%. Interplay between the factors affecting it is complex. For example, in assessing intervention designs, modelling has suggested that mandated isolation, if it leads to substantially reduced uptake of testing, could overall have an undesirable impact on the pandemic [45] (although this went on to become policy). The communication of personalised risks relating to SARS-CoV-2 is also challenging, with research suggesting that absolute likelihoods of risks may be considered less helpful than describing major risk factors for different ‘personas’ [18]. This may signify different risk demographics, another factor of interest to modelling.

It is important to find TTI strategies that are effective but which are also acceptable and feasible to adhere to by members of the public, in all of their diversity. TTI policy design choices that seem optimal when assuming 100% adherence might not actually be the most effective policies in practice. Conducting these interviews has helped us to understand what aspects of the TTI policy different people might find most costly and help us to identify alternative strategies, which could then be assessed within a modelling framework.

## 6 Strengths and limitations

This is a qualitative interview study and findings cannot be generalised. Interviews took place during a policy change (19 July 2021), so the findings may be over-representing aspects of the interview context. Whilst efforts were made to recruit a diversity of views, participants were necessarily those who chose to take part in this study. As noted by [4], it may be that disengaged views are not adequately represented.

We did not capture quotes from interview participants for increased confidentiality, so these are not used in the reporting of the study. The participant sample was partially convenience based and reporting views on hypothetical or observed situations.

This study has provided insights into public interactions and perceptions of TTI systems in the UK, which can be used towards improving policy effectiveness. We have also highlighted a range of ways in which this information has bearing on the methods and interpretation of TTI modelling in collaboration with a group of modellers who have been heavily involved in UK government scientific advice on this topic through 2020-2021. It represents a step towards public involvement and engagement with rapid pandemic response infectious disease modelling research, rather than being a proposed framework or example of full co-production between researchers and members of the public. We experienced challenges in identifying methods to represent and communicate model processes and assumptions in a form that members of the public could interact with. Further, our limited time frame and the technical complexity meant we were not able to conduct a systematic and comprehensive review or assessment of modelling assumptions. These are topics for further work.

## 7 Conclusion

Epidemics are dynamic situations and a process whereby the population learns how to “live with the virus” (immunologically, epidemiologically, socially and personally), so there is no perfect point in time from which to generalise. This study took place during a period of constraint relaxation, but as noted in the Discussion some of the findings match well to prior research conducted a year previously. Our research suggests the usefulness of engagement with the Public in order to test, validate or explore modelling assumptions.

In this interview study, we identified key themes relating to TTI in the UK: Education and guidance, trust, practicality, mental health, and perceived risk, together with attitudes about the future and recommendations for TTI engagement and control of SARS-CoV-2. These themes, with more granular insights such as the challenges of testing children and the clustering of culture-related behaviours, provided stimuli for TTI modellers to reflect on their assumptions and decisions.

Our study points to the importance of engaging with the public at all stages of an infectious disease modelling response project. The value of cross-disciplinary input is recognised and reflected in the composition of pandemic government scientific advisory groups [53], but the nature of our public interview findings have implications for the setting and framing of research questions, and fundamental choices of model structure. These issues go to the heart of how research agendas regarding pandemic control intervention choice and design decisions are set, though a full discussion is beyond the scope of this paper. During the COVID-19 pandemic in the UK, modellers have been engaged in responding to sometimes very specific questions posed by decision-makers to the SPI-M scientific advisory committee, though sometimes modellers have themselves initiated the investigation of particular interventions or forms by which they are implemented and brought them to the committee [53]. However, even within a pre-specified request, with reflection against what can be learned from public engagement with policies in practice, we can see that there might be room still for decisions to have been made unawares or implicitly to make decisions or frame analyses in such a way as to obscure or exclude important considerations or equity issues [71]. Especially in the rapidly shifting and urgent epidemiological and policy context of a pandemic, regular cycles of feedback and guidance would be of benefit. For policy-makers, this study points towards the importance of social coherence and common perceptions and common aims for epidemic management. The advice to ensure full participation of the public and stakeholders throughout the life-cycle of an academic public health project is not new [64] but there is relatively little literature specifically discussing the barriers and facilitators to doing this for infectious disease modellers. The literature that there is highlights major challenges and the need for longer-term public-research collaboration [54]. As conclusions are drawn about the necessary set of scientific advisory and pandemic research tools, we argue for the further development of public participation processes for pandemic response infectious disease modelling.

## Data Availability

All data produced are available online at https://doi.org/10.6084/m9.figshare.15067119

## Data accessibility

Data and supporting resources developed and used in the study are shared via a publicly accessible repository [17].

## Authors’ contributions

G.M. and R.S. carried out the study and analysed results. G.M, R.S and E.F drafted the manuscript. G.M, R.S, C.J, L.Y and E.F were involved in the conceptualisation and study design. M.E.P.S, M.F, T.H, L.P, E.L.D, G.F.M, B.J.Q, L.D and E.F provided insights in Section 5. All authors were involved in revising of the manuscript and approved the final manuscript.

## Acknowledgment

We thank the project team of ‘An Analytical framework for Test, Trace and Isolate in the UK’ for input into the study design, and Adam Kucharski, who attended the workshop but was not able to collaborate on the writing of this paper.

